# Social and emotional characteristics of girls and young women with *DDX3X*-linked intellectual disability: A descriptive and comparative study

**DOI:** 10.1101/2021.09.23.21264040

**Authors:** Elise Ng-Cordell, Anna Kolesnik-Taylor, Sinéad O’Brien, Duncan Astle, Gaia Scerif, Kate Baker

## Abstract

**Background:** *DDX3X* variants are a common cause of intellectual disability (ID) in females. Previous studies have reported high rates of autism spectrum disorder and emotional-behavioural difficulties within this group. However, no study has compared social and emotional characteristics in individuals with *DDX3X* variants to individuals with other monogenic causes of ID.

**Methods:** Twenty-three females with pathogenic or likely pathogenic *DDX3X* variants were recruited via UK regional genetics centres, genetic research cohorts, and family support groups. Twenty-three females with ID and variants in other genes were recruited via the same routes. Phenotypic data were collected through semi-structured interviews and standardised questionnaires.

**Results:** In line with previous research, we found a wide range of adaptive, social and emotional function within the *DDX3X* group. Autism characteristics assessed via the Social Responsiveness Scale (SRS) did not differ between *DDX3X* and comparison groups, while levels of anxiety and self-injurious behaviour (SIB) scores assessed via the Developmental Behaviour Checklist (DBC) were significantly higher in the *DDX3X* group. Autistic characteristics, anxiety and SIB scores were positively correlated within the *DDX3X* group. Individuals with *DDX3X* missense variants had poorer adaptive abilities than those with protein truncating variants as previously reported, but *DDX3X* variant type did not significantly predict emotional and behavioural characteristics.

**Conclusions:** We provide quantitative evidence that overall incidence of autistic characteristics is not higher amongst girls and women with *DDX3X* variants than expected for ID. However, the *DDX3X* group demonstrates more SIBs and anxiety symptoms than expected, with close relationships between SIBs and anxiety, and between anxiety and autistic characteristics. Future work is warranted to explore the multilevel mechanisms contributing to social and emotional development in individuals with *DDX3X* variants, and consider whether these mechanisms are specific to this genetic diagnosis or shared with a subset of people with ID due to other causes.

## Background

At least 60% of individuals with intellectual disability (ID) have an underlying genetic diagnosis (1). With advances in next generation sequencing, it is increasingly possible to diagnose genetic causes of ID, representing an opportunity to study aetiological factors influencing development and lifespan experiences among individuals with ID. With ever increasing numbers of rare and ultra-rare genetic diagnoses associated with ID, it is especially important to place the phenotypic spectrum of each diagnosis in the context of expectations for ID in general, and to identify factors associated with variability within genetic diagnosis groups. Comparative phenotyping studies can thus highlight whether features occur at elevated rates, or whether they occur in line with expectation for ID more generally (2). In addition, studying relationships between phenotypic dimensions can be a starting point towards understanding mechanisms contributing to mental health across syndromes.

*DDX3X* loss-of-function variants are one of the more common single-gene causes of ID, affecting an estimated 1-3% of females with unexplained ID (3, 4). As availability of genetic testing increases, numbers of individuals diagnosed with *DDX3X* variants are likely to rise considerably. The *DDX3X* gene is located at Xp11.4, and encodes an ATP-dependent DEAD-box RNA helicase (5, 6) involved in multiple aspects of RNA metabolism including splicing and transport (7), mitochondrial DNA production (4), and the Wnt/Beta-catenin signalling pathway (3). Animal models have shown that *DDX3X* is highly expressed in radial glial progenitors and neurons, and promotes cortical development by regulating neurogenesis and migration (8, 9). Neuroimaging has highlighted diverse neuroradiological features (e.g. corpus callosum abnormalities, ventricular enlargement, cortical dysplasia, polymicrogyria), reflecting the regulatory functions of *DDX3X* during multiple phases of brain development (9-11). Hence, there are diverse molecular, cellular and neural systems mechanisms by which pathogenic *DDX3X* variants may influence emergent cognition and behaviour.

Previous studies of individuals with *DDX3X* variants have primarily relied upon retrospective collation of clinical records, and have described a set of variable and complex clinical characteristics. Common physical features include cardiac and respiratory problems (e.g. sleep apnoea, congenital heart disease) and musculoskeletal disorders (e.g. joint hypermobility, scoliosis), while a range of neurodevelopmental characteristics have been reported, encompassing global cognitive impairments from mild to severe ID, autism spectrum disorder (autism), hyperactivity, aggression, and motor stereotypies (3, 4, 9-13).

Detailing the social and emotional characteristics of individuals with *DDX3X* variants, including factors underlying variation in outcomes, is important for understanding the support needs of diagnosed individuals as well as their families and caregivers (14, 15). Lennox et al (9) reported that among 53 individuals with *DDX3X* variants, average adaptive ability fell within the moderate ID range, while autism characteristics assessed via the Social Responsiveness Scale (16) and the Social Communication Questionnaire (17) were moderately elevated above general population norms (not stratified for ID). Further, they reported that missense variants were associated with more severe clinical outcomes across neuroanatomical, neurological, adaptive and behavioural domains. Elsewhere, Tang et al (18) characterized behavioural profiles of 15 individuals (14 females) with *DDX3X* variants via systematic post-diagnostic assessments of intellectual and adaptive functioning, sensory processing, autism and behavioural comorbidities. Based on diagnostic clinical assessments, 80% of individuals with *DDX3X* variants met diagnostic criteria for ID, 60% for autism, and 53% for attention deficit / hyperactivity disorder (ADHD). Comparing scores on standardized questionnaire measures to population norms revealed high rates of sensory processing abnormalities such as sensory-seeking behaviours and sensory hypo-reactivity. In addition, 14% of individuals reported elevated anxiety above a likely clinically-relevant level. Comparing behavioural profiles across variant types revealed that missense variants and in-frame deletions were associated with poorer language, motor and adaptive outcomes compared to protein-truncating variants (18). Further, a smaller proportion of individuals with protein-truncating variants met diagnostic criteria for autism, although this difference was not significant and may reflect expected differences between groups with different adaptive abilities.

In summary, research to date points towards prevalent and diverse social and emotional difficulties in individuals with *DDX3X* variants. Moreover, variant type appears to be one aetiological factor underlying phenotypic variability. However, the distinctiveness of these characteristics from other causes of ID in females with a similar range of global adaptive severity, and relationships between adaptive-social-emotional dimensions, have not been established. Therefore, in the current study, we aimed to 1) describe the clinical, social, and emotional characteristics of girls and women with *DDX3X* variants, 2) identify social and emotional features that differentiate females with *DDX3X* variants from females with other monogenic causes of ID, and 3) explore predictors (variant type, ID severity, autistic characteristics) of social and emotional outcomes among individuals with *DDX3X* variants.

## Methods

### Participants and recruitment

Individuals who had previously received a diagnosis of a pathogenic variant in *DDX3X* were identified via clinical genetics services across the UK, the IMAGINE-ID (Intellectual Disability & Mental Health: Assessing the Genomic Impact on Neurodevelopment) Study (https://imagine-id.org/), or the *DDX3X* Support UK Group (https://ddx3xsupportuk.co.uk/). Clinicians were given recruitment packs to distribute to family members of each potential participant. Members of the IMAGINE-ID Study and the *DDX3X* Support UK Group were provided with information and contact details for the study. The families of 23 individuals volunteered to take part (n = 8 via clinical genetics services, n = 6 via the IMAGINE-ID Study, and n = 9 via the *DDX3X* Support UK Group). All 23 participants had been diagnosed with *DDX3X* variants through research or clinical pathways via whole exome sequencing or gene panel testing. Of the 20 participants for whom we had variant details, 7 had missense variants and 13 had protein truncating variants (protein truncating variants included frameshift, nonsense and splice site variants; Supplementary Table 2). The same procedures were followed to recruit a comparison group of 23 girls and young women with ID of monogenic origin (ID-comparison group), represented by pathogenic variants in 11 different genes (Supplementary Table 1).

### Phenotyping assessments

Assessments were carried out at participants’ homes for 12 families. For the remaining 11 families, assessments were conducted via telephone or video conference. We conducted a structured medical history interview (MHI), followed by the Vineland Adaptive Behaviour Scales (VABS): we administered either the second edition, interview form (19; n = 12), or the third edition, parent/caregiver form (20; n = 13). In addition, parents completed the Social Responsiveness Scale, second edition (SRS; 16) and the Developmental Behaviour Checklist – second edition (DBC; 21). Following the above assessments, we administered the Autism Diagnostic Interview – Revised (ADI-R; 22); however, a shortened timeline due to the COVID-19 pandemic meant that we were only able to collect ADI-R data from a subset of seven participants in the *DDX3X* group, and no participants in the comparison group.

### Analysis Plan

Raw subscale and total scores from questionnaire data were converted to standardised scores based on published normative data.

#### Aim 1

We described the clinical, adaptive, social, and emotional characteristics of the *DDX3X* participants, by summarizing reports from the MHI, VABS, SRS, ADI-R, and DBC.

#### Aim 2

To identify adaptive, social, or emotional characteristics that might differentiate *DDX3X* variants from other causes of ID in females, we compared the *DDX3X* group to the ID-comparison group on the VABS, SRS, and DBC. Independent samples t-tests and non-parametric tests (Mann-Whitney U) were used to compare scores on the SRS and DBC. To compare scores on the VABS, we conducted univariate analyses, controlling for VABS form (i.e. second or third edition).

#### Aim 3

We ran pairwise correlations, to explore potentially distinct relationships between age, ID severity (VABS Composite), autistic characteristics (SRS Total), anxiety (DBC Anxiety), and self-injurious behaviours (SIBs; calculated by summing scores of three DBC self-injury items) within the *DDX3X* and ID Comparison groups separately. To follow up, we ran general linear models within the *DDX3X* group, to investigate whether variant type (missense vs. protein truncating), ID severity, or autistic characteristics predicted levels of anxiety and SIBs.

## Results

### Aim 1: Describing the *DDX3X* group

#### Clinical and developmental characteristics

Clinical and developmental characteristics, taken from the MHI, are summarized in Table 1. Impairments affecting nearly all participants (>90%) included delays in achieving communication and motor milestones, as well as gastrointestinal problems (most commonly feeding difficulties in infancy, and ongoing, chronic constipation). Common impairments (affecting >50% of participants) included complications during pregnancy and childbirth, hypotonia during infancy, hearing and vision impairments, sleeping difficulties, movement disorders, musculoskeletal abnormalities, and self-injurious behaviours. Characteristics that were observed in <50% of participants included sensory hypo/hyper-sensitivities (including a high pain threshold or sensory-seeking behaviours), respiratory and cardiological issues, skin conditions, minor congenital abnormalities, and epilepsy.

**Table 1.** Clinical phenotypes summary.

| Clinical feature | Frequency of feature (n/23) | Frequency of feature (%) | Subtypes and frequencies (n) |
| --- | --- | --- | --- |
| **Gastrointestinal** | 22 | 96 | *Infancy:* Latching / sucking difficulties (14), Reflux (9), Choking (3)<br>*Current / childhood:* Constipation / Immotile gut (8), Overeating (3), Overfills mouth (2), Incontinence (1) Cyclical vomiting (1) |
| **Delayed communication milestones** | 21 | 91 | Mild = using words and phrases (11), Moderate = using single words only (5), Severe = not using any words (4), Unable to classify as under age 5 years or insufficient information (1) |
| **Delayed motor milestones** | 21 | 91 | Mild = walked by 3 years (15), Moderate = walked by 5 years (6) |
| **Prenatal, Perinatal, and Neonatal complications** | 15 | 65 | *Prenatal:* Ultrasound abnormalities (5), Small foetus (3), Preeclampsia (1)<br>*Perinatal:* Foetal heart rate fluctuation during delivery (4), Breech positioning (3), Umbilical cord abnormalities (3),<br>*Neonatal:* Excessive crying (4), Jaundice (1), Meningitis (1) |
| **Hearing / Auditory** | 14 | 61 | Recurrent ear infections in infancy / early childhood, often leading to partial hearing loss (8), Hypersensitivity to sounds (8) |
| **Ophthalmic** | 13 | 57 | Far-sightedness / hyperopia (3), Cortical visual impairment (3), Squint (2), Nystagmus (1), Astigmatism (1), Abnormal pupil dilation (1), Poor depth perception (1), Blocked tear ducts (1), Eye tracking difficulties (1) |
| **Infantile hypotonia** | 13 | 57 | Parents usually noticed poor head control and ability to sit up during first year |
| **Movement problems** | 13 | 57 | Poor balance and coordination (10), Stiff gait (7), Stereotypies (3), Tremor (2), Ataxia (1), Dystonia (1) |
| **Sleep difficulties** | 13 | 57 | *Infancy:* Hypersomnia (4), Hyposomnia (2)<br>*Current / childhood:* difficulties falling and staying asleep, early waking (9) |
| **Self-injury** | 13 | 57 | Hitting / biting (13), Skin scratching / picking (13), Head banging (11), Hair pulling (2) |
| **Musculoskeletal** | 13 | 57 | Joint Hypermobility (7), Hypotonia in limbs (4), Scoliosis (4), Inward-turning feet (2), Bone deficiency (1) |
| **Sensory** | 10 | 43 | Parents often reported a combination of sensory-seeking tendencies and sensory aversions, as well as a high pain tolerance. |
| **Respiratory / cardiology** | 10 | 43 | Frequent colds / respiratory infections (5), Heart murmur (4), Hay fever (3), Unspecified heart problems (2), Narrow heart valve (1), Asthma (1), Sleep apnoea (1) |
| **Skin conditions** | 8 | 35 | Eczema (6), Impetigo (1), Palmoplantar keratosis (1) |
| **Minor congenital abnormalities** | 5 | 22 | Inguinal hernia (2), Finger abnormalities (2), Adrenal hyperplasia (1), Cleft palate (1) |
| **Epilepsy** | 6 diagnosed | 26 | *Diagnosed:* Absence seizures (2), Generalized seizures (1), Infantile spasms (1), Reflux anoxic seizures (1), Unspecified (3)<br>*Suspected:* Absences / isolated seizures in past (8) |
|  | 8 suspected | 35 |  |
| **Other** | 7 | 30 | Allergic reactions / oedema (3), Recurrent UTIs (2), Kidney Stones (1), Diabetes (1), Poor circulation in extremities (1), stereotypic laughter (1) |

#### Adaptive functioning

Adaptive functions, social, and emotional characteristics are summarized and displayed in Table 2. Global adaptive ability was measured using the VABS composite score. Average composite score was 53.09 (SD = 14.42; moderate ID range), although level of impairment ranged from borderline to profound (borderline n= 2, mild n= 9, moderate n= 8, severe n= 3, profound n= 1). Following published guidelines (15, 16), we examined domain-level scores (in Communication, Daily Living Skills, and Socialisation domains) to identify areas of relative strength or weakness. Communication (receptive, expressive, and writing skills) was a relative weakness for seven participants (and a relative strength for one). Daily Living Skills (personal, domestic, and community skills) were a relative weakness for seven participants (and a relative strength for one), while Socialisation (interpersonal relations, play and leisure, and coping skills) was an area of relative strength for eight individuals (Socialisation was not an area of weakness for any participants).

**Table 2.** Adaptive, Emotional and behavioural characteristics.

|  | DDX3X |  |  | ID-comparison |  |  | Group difference |
| --- | --- | --- | --- | --- | --- | --- | --- |
|  | N | Mean (SD) | Range | N | Mean (SD) | Range | DDX3X – ID-Comparison |
| **Age** | 23 |  |  |  |  |  |  |
|  |  | 12.87 (5.50) | 3-22 | 23 | 13.20 (5.15) | 6.72-26.28 | t(44) = -.21, p = .835, d = .062 |
| **VABS** | 23 |  |  | 23 |  |  |  |
| ABC | | 53.09 (14.42) | 20-74 | | 55.61 (11.78) | 34-79 | F = .115, p = .736, $\eta^2$ = .003 |
| Communication | | 50.26 (17.41) | 20-79 | | 57.65 (14.25) | 33-89 | F = .002, p = .966, $\eta^2$ = .000 |
| Daily Living | | 48.65 (16.25) | 20-73 | | 54.74 (14.99) | 30-83 | F = 1.017, p = .319, $\eta^2$ = .023 |
| Socialization | | 57.48 (15.17) | 20-78 | | 59.22 (11.03) | 42-88 | F = .006, p = .507, $\eta^2$ = .000 |
| Motor | 19 | 67.79 (11.87) | 45-97 | | 66.26 (17.75) | 28-121 | F = .185, p = .669, $\eta^2$ = .005 |
| **DBC2** | 21 |  |  | 23 |  |  |  |
| Total |  | 58.86 (16.03) | 36-106 |  | 53.57 (12.11) | 34-83 | t(42) = 1.24, p = .22, d = .38 |
| Disruptive |  | 54.14 (16.11) | 38-112 |  | 50.22 (11.33) | 35-77 | t(42) = .94, p = .35, d = .28 |
| Self-absorbed |  | 63.90 (17.66) | 39-104 |  | 56.22 (12.96) | 37-83 | t(42) = 1.66, p = .11, d = .50 |
| Communication |  | 59.29 (19.46) | 42-132 |  | 55.78 (10.84) | 36-81 | t(42) = .75, p = .46, d = .23 |
| Anxiety |  | 55.57 (10.57) | 38-83 |  | 50.04 (12.25) | 36-77 | U = 147.50, Z = 2.21, p = .03 |
| Social relations |  | 51.67 (11.70) | 36-76 |  | 51.04 (10.31) | 38-73 | t(42) = .19, p = .85, d = .06 |
| **SRS** | 21 |  |  | 22 |  |  |  |
| Total |  | 77.38 (14.08) | 46-98 |  | 76.82 (14.54) | 49-96 | t(41) = .13, p = .90, d = .04 |
| Awareness |  | 73.52 (11.85) | 50-93 |  | 76.09 (14.53) | 49-96 | U = 193.00, Z = -.93, p = .35 |
| Cognition |  | 74.43 (11.08) | 51-92 |  | 73.59 (14.88) | 41-96 | t(41) = .21, p = .84, d = .06 |
| Communication |  | 75.86 (13.60) | 42-96 |  | 74.32 (12.85) | 50-91 | t(41) = .38, p = .71, d = .12 |
| Motivation |  | 66.29 (14.00) | 44-95 |  | 65.59 (13.58) | 41-89 | t(41) = .17, p = .87, d = .05 |
| RRBs |  | 80.57 (17.18) | 48-108 |  | 78.59 (16.10) | 48-106 | t(41) = .38, p = .70, d = .12 |

#### Social and autistic characteristics

The SRS was used to measure autistic characteristics. Total T-scores ranged from 46 to 98, with an average score of 78.00 (SD = 14.15; a T-score of 75 indicates highly elevated symptoms that are characteristic of autism). Four participants had scores that fell in the normal to mildly elevated range, four had scores in the moderately elevated range, and twelve had scores in the highly elevated range. Scores across subscales varied, with average levels of Social Awareness, Social Cognition, and Social Motivation falling within the mild-moderately elevated range, and average levels of Social Communication and Restricted, Repetitive Behaviours falling within the highly elevated range.

To explore autistic characteristics of girls with *DDX3X* variants in more depth, we completed the ADI-R with the parents of seven participants. Based on published criteria (22), one participant met the current threshold for autism in all three domains (Socialisation, Language and Communication, Restricted and Repetitive Behaviour), while three met or exceeded cut-off scores on two of the three domains. Individual item scores on the ADI ranged from 0-3, with higher scores indicating more autism-like characteristics. To assess whether there is consistency in specific autistic characteristics within the DDX3X group, we reviewed average scores on each ADI item (supplementary Figure 1 – current behaviours, supplementary Figure 2 - scores at age 5, or the “most abnormal” time period). Items that had the highest endorsement (mean score > 1.5) related to difficulties with friendship and sharing (from the Social Development Domain), verbal intonation, reciprocal conversation, and imaginative play (from the Language and Communication Domain), and auditory sensitivity (from the Interests and Behaviours Domain). Conversely, items that were seldom endorsed (mean score < 0.5) related to making social overtures, offering comfort to others, and responding to other children (Social Development Domain), communicative speech, idiosyncratic use of language, and non-verbal gestures (Language and Communication Domain), and preoccupations, compulsions, and inflexibility (Interests and Behaviours Domain).

#### Emotional and behavioural characteristics

The DBC was used to assess emotional and behavioural difficulties. DBC scores varied widely in the *DDX3X* group, with total problem behaviour T-scores (stratified by ID severity) ranging from 36 to 106. Thirteen of the twenty-one participants who completed the DBC had total scores that exceeded the clinical cut-off, indicating severe concerns across emotional and behavioural domains. Among the subscales, Anxiety was most commonly rated as an area of Serious concern (16/21 participants). Frequently endorsed anxiety-related behaviours included shyness and social withdrawal, marked worries about routine, and unusual or excessive fears of sounds or objects (including fire alarms, babies crying, hand driers, sneezing, fireworks, onions, or butterflies - for one participant, fears of animals with stings had transformed into an obsession). The Self-Absorbed subscale, which largely captures autism-like behaviours, was also commonly rated as an area of Serious concern (15/21 participants). Items frequently endorsed on this scale related to arranging objects in a strict order and sensory-seeking behaviours.

As noted above, thirteen participants displayed self-injurious behaviours (SIBs). During the MHI, parents reported that their daughters responded to stressful or frustrating situations by pulling their hair, biting their hands or knees, banging their head, or throwing themselves onto the floor. To further investigate the types and frequencies of SIBs demonstrated by participants, we also examined three items of the DBC pertaining to these behaviours: 11 participants banged their heads, 13 participants hit or bit themselves, and 13 participants picked or scratched their skin.

### Aim 2: Comparing adaptive, social and emotional characteristics in the *DDX3X* and ID-comparison groups

Next, we explored whether any adaptive, social or emotional characteristics consistently differentiated the *DDX3X* group from girls and women with ID, by comparing the *DDX3X* and ID-comparison groups using scores from the VABS, DBC, and SRS (Table 2). There were no significant differences between the two groups on the VABS or the SRS. On the DBC, the *DDX3X* group had higher scores on the Disruptive, Self-Absorbed, Communication, and Anxiety subscales, with effect sizes ranging from small to medium (Figure 1). Furthermore, the group difference on the Anxiety subscale was significant (Mann-Whitney U test statistic = 147.50, *p* = .027).

**Figure 1.**
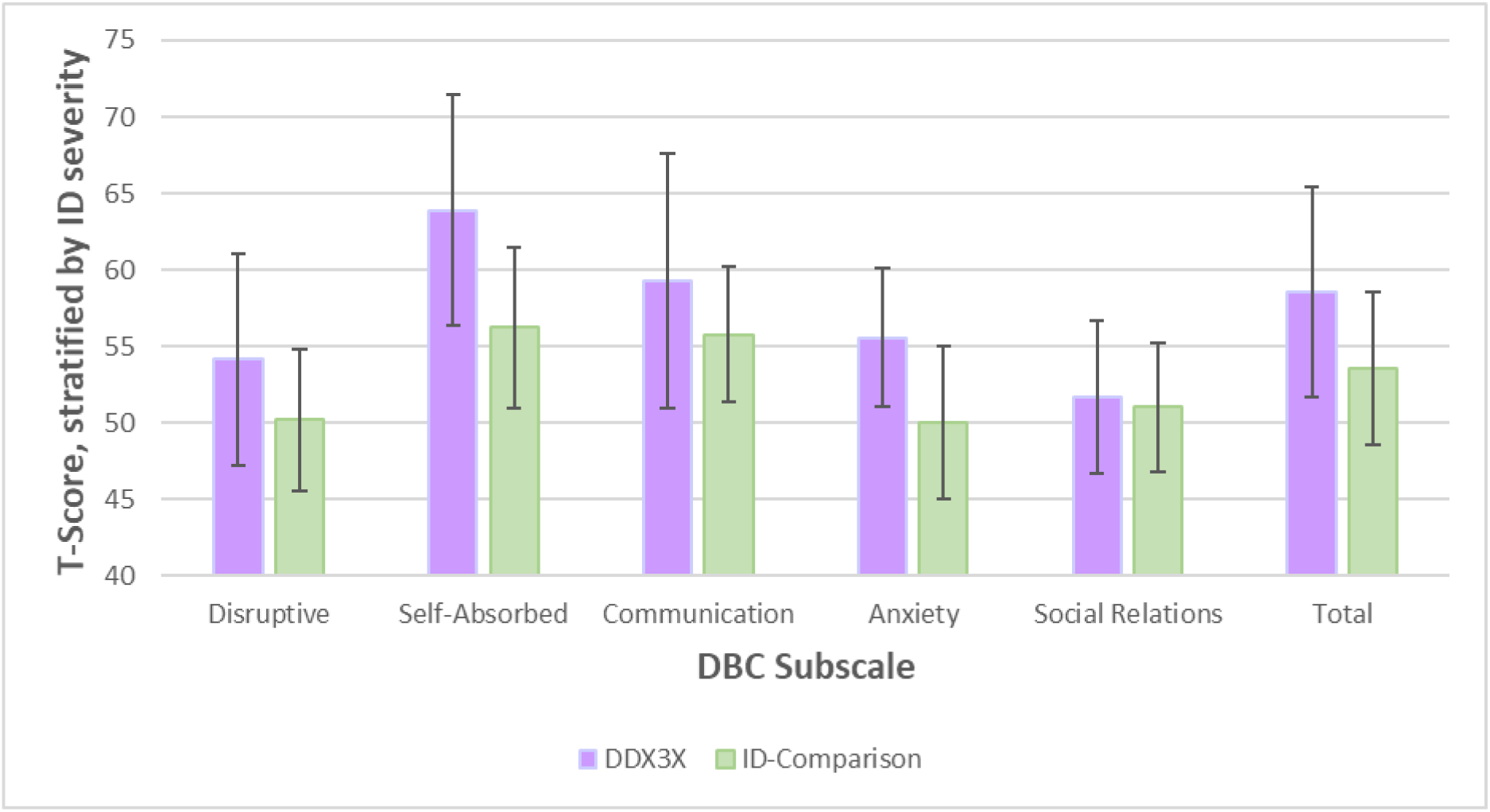
Emotional and behavioural characteristics across the DDX3X and ID-Comparison groups. *Note*. Error bars represent 95% confidence intervals

To follow-up on parent reports of SIBs in the *DDX3X* group, we also explored whether rates of SIBs (as measured by three items on the DBC) were elevated in this group relative to the ID-comparison group. A higher proportion of participants in the *DDX3X* group showed head banging (χ^2^ = 5.98, p = .014) and self-hitting/biting (χ^2^ = 4.39, *p* = .036). Skin picking/scratching was more common in the *DDX3X* group, but not significantly different from the comparison group (χ^2^ = 3.24, *p* = .072).

### Aim 3: Within-group predictors of social and emotional outcomes

Our third aim was to examine relationships between age, ID severity (VABS Composite), autistic characteristics (SRS Total), anxiety (DBC Anxiety), and SIBs (calculated by summing scores on the three DBC self-injury items) within the *DDX3X* and ID-Comparison groups separately. Bivariate correlations are displayed in Figure 2.

**Figure 2.**
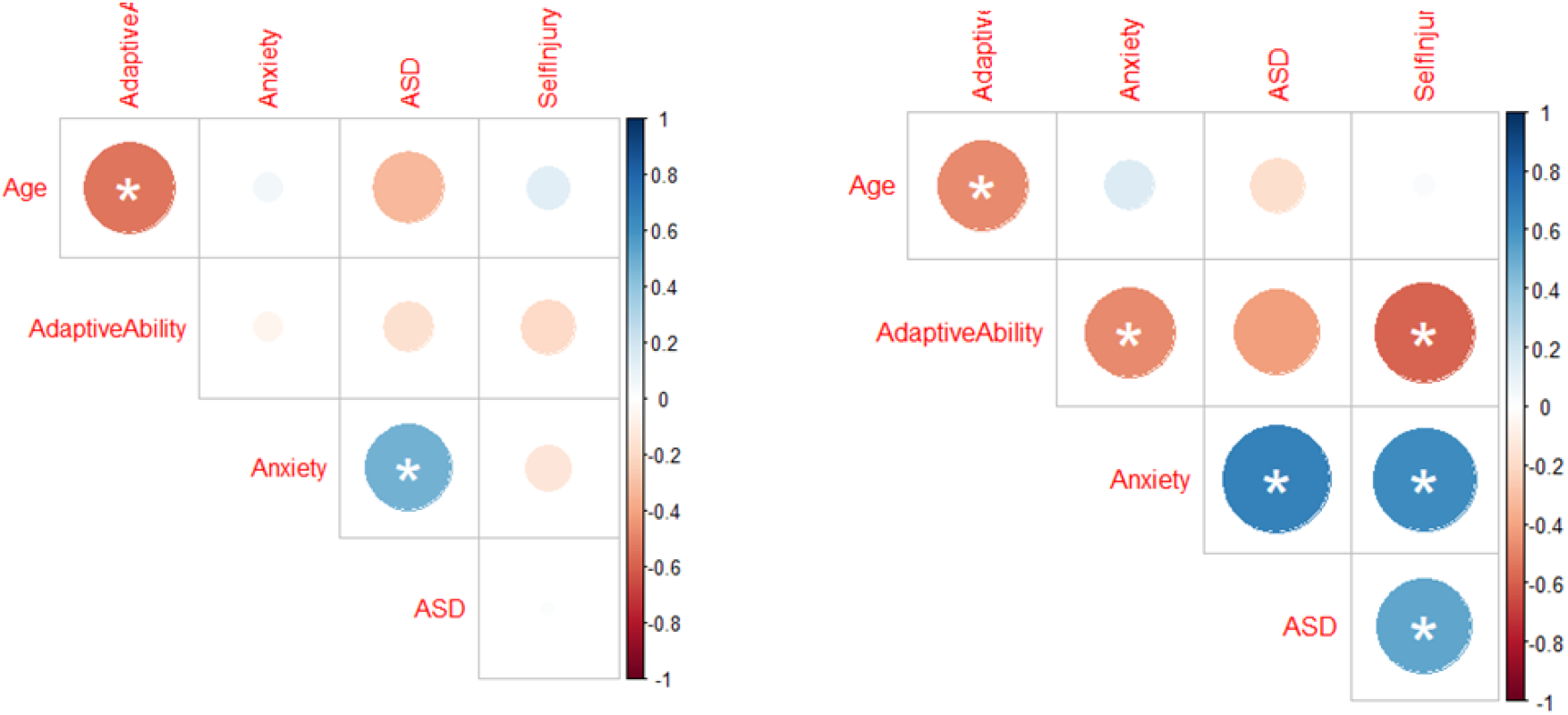
Correlation plots for the ID-comparison (left) and DDX3X (right) groups.

In both groups, older age was associated with lower adaptive ability (*DDX3X*: *r* = -.47, *p* = .02; ID-comparison: *r* = -.40, *p* = .06), and higher anxiety was associated with elevated autism characteristics (*DDX3X*: *r* = .68, *p* = .001; ID-comparison: *r* = .43, *p* = .05). At the same time, several significant correlations emerged in the *DDX3X* group only. Within the *DDX3X* group, higher anxiety was associated with lower adaptive ability (*r* = -.48, *p* = .03) and greater self-injurious behaviours (*r* = .62, *p* = .003). In addition, self-injurious behaviours were associated with elevated autism characteristics (*r* = .53, *p* = .02).

Finally, we ran two linear models to explore factors contributing to variation in commonly endorsed emotional characteristics (anxiety and SIBs) within the *DDX3X* group. In both models, we included age and variant type (missense versus protein-truncating) as predictors. As has been reported previously (18), participants with missense variants had poorer adaptive abilities than those with protein truncating variants (missense: mean VABS Composite = 43.71, SD = 12.67; protein truncating: mean VABS Composite = 58.77, SD = 14.56); we therefore controlled for adaptive ability in our models. Informed by our earlier correlation analyses, we included autistic characteristics in the model predicting levels of anxiety, and anxiety in the model predicting levels of SIBs. Linear models are presented in Table 3. In the first model, anxiety was associated with autism characteristics (β = .70, *t* = 3.22, *p* = .007) such that higher levels of autistic characteristics predicted higher levels of anxiety. In the second model, SIBs were associated with anxiety (β = .54, *t* = 2.67, *p* = .019), such that higher levels of anxiety predicted more SIBs. Age, adaptive ability and variant type did not significantly predict anxiety or SIBs.

**Table 3.**
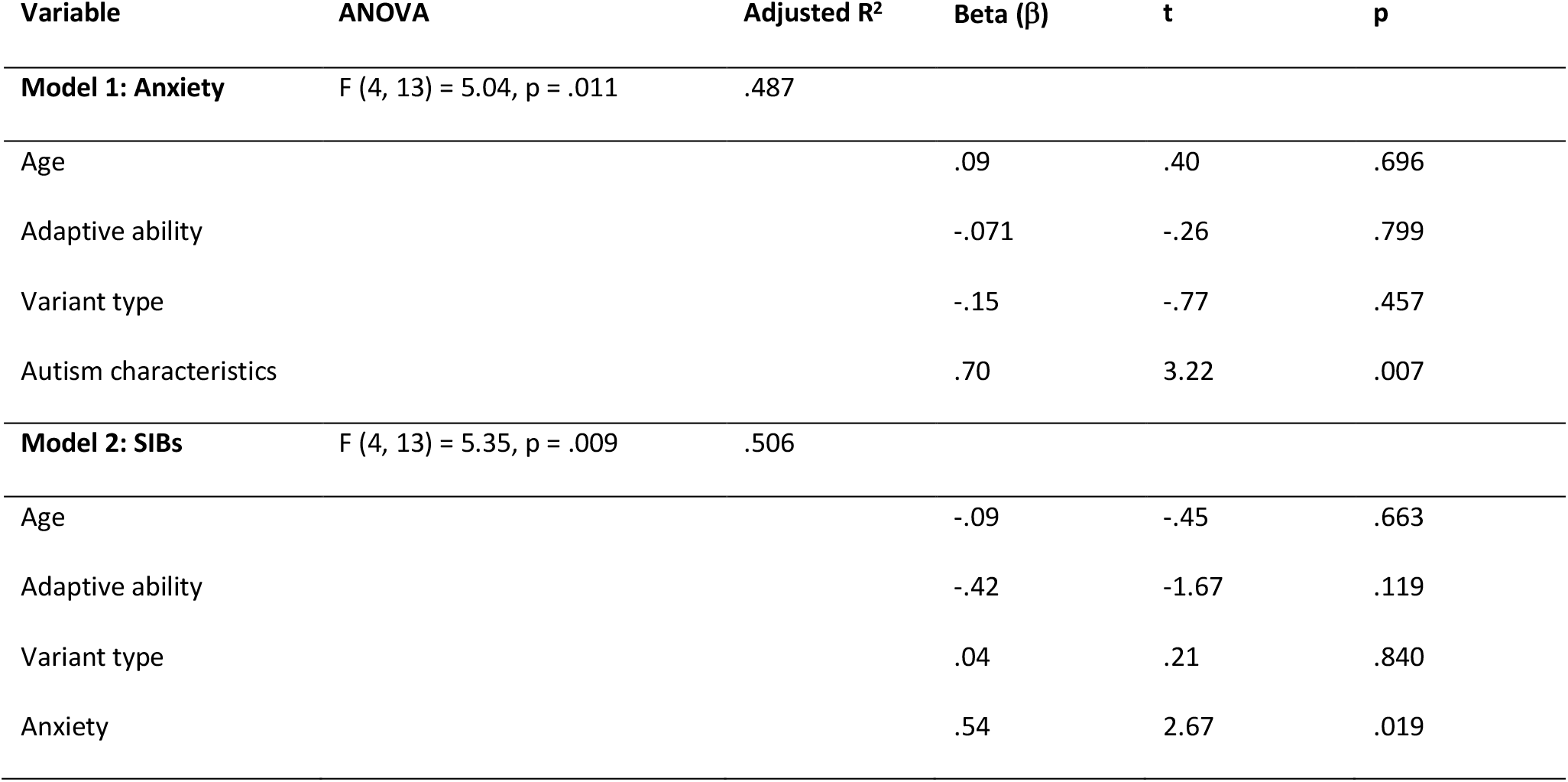
Linear regression models.

## Discussion

In this study, we build on previous characterisations of girls and women with *DDX3X* variants, in order to enhance prognostication, management and support for families and individuals with this diagnosis. We focus on social and emotional development, as these areas of need have been highlighted in published literature (9, 18), but not yet deeply explored. Our primary objective was to determine whether the neurodevelopmental characteristics of this highly variable disorder are as expected for girls and young women with ID of any cause, or are in any respects specific to *DDX3X*. Below, we highlight novel observations not previously reported in the literature on *DDX3X*, discuss evidence for specificity of neurodevelopmental vulnerabilities, and consider potential explanations for the co-occurrence of specific characteristics within this group.

By conducting medical history interviews with a relatively large group of parents and carers, we identified several common areas of difficulty which have not been emphasised in previous reports. These included notably high frequency of early life feeding difficulties and sensory difficulties, which are important for understanding the experiences of parenting a young child with *DDX3X*, and are potentially relevant to emerging cognitive and social-emotional traits. Of particular note, a very high proportion of parents reported symptoms of self-injurious behaviours in their daughters, which have not been reported previously in association with this genetic diagnosis. Furthermore, many parents described their daughter as having a strong desire to make friends, yet lacking the skills to develop and sustain friendships, and tending towards heightened self-consciousness in social situations. Importantly, we observed these social and emotional challenges across the wide spectrum of ages and adaptive abilities in the *DDX3X* group. In addition to the diversity of challenges reported, parents described many of their daughters’ strengths and qualities, including caring and friendly personalities, humour and laughter, love of dancing and swimming, and willingness to persevere in the face of challenges.

Motivated by information elicited via these semi-structured interviews, we proceeded to systematically evaluate the social and emotional characteristics of the *DDX3X* group using a variety of questionnaire and interview methods. SRS scores indicated that many girls with *DDX3X*-linked ID exhibit autistic characteristics, with the range of SRS scores aligning with previous reports (9, 18). However, SRS scores (total and subscales) did not differ between the DDX3X and ID-comparison groups, suggesting that overall likelihood of autistic characteristics as assessed by this tool is not a discriminating feature of *DDX3X*. Conducting the ADI with a subset of the *DDX3X* group highlighted a focused set of social vulnerabilities, cutting across traditional autism domains. Previous research in girls and women with fragile X syndrome (FXS) suggests a similar profile of social difficulties, characterized by preserved empathy alongside broader social communication challenges and repetitive behaviours (23, 24). In the context of this literature, our findings suggest that the social vulnerabilities observed in association with *DDX3X* variants reflect autistic characteristics shared amongst females with monogenic ID; however, further work is required to confirm this.

In contrast, anxiety scores significantly differentiated the *DDX3X* group from the comparison group. Rates of reported anxiety, which was a concern for a substantial majority of participants in our study, are considerably higher than previously reported by Tang et al (18). One possible explanation for this discrepancy is that the Child Behaviour Checklist (25) used in previous studies may be less sensitive than the DBC to anxiety in young people with autism and/or ID (26). Another striking and previously unreported feature of the *DDX3X* group was the high incidence of self-injurious behaviours (SIBs), reported as a current or recent distressing problem for more than half of participants, and significantly more common amongst the *DDX3X* group than the comparison group. In summary, anxieties and SIBs can be anticipated for this group, signalling elevated need for clinical psychology support during childhood and adolescence.

These findings suggest that while the *DDX3X* gene has received a great deal of interest for its association with autism, it is equally important to recognise and address co-occurring anxiety in this group. Indeed, features of anxiety and autism often overlap (e.g. sensory sensitivities, intolerance of uncertainty, social avoidance), such that anxiety symptoms can often be misattributed to autism in clinical practice (see 27 for review). The presentation and needs of autistic girls and women with ID are often under-recognized and unmet, adding further complexity (28, 29). To address this, we evaluated the associations between autistic characteristics and anxiety within the DDX3X group, finding that they were closely associated and not simply a reflection of ID severity and adaptive function.

Turning again to the FXS literature, anxiety among girls and women with FXS can, in part, be attributed to autism-related social difficulties (30). Mechanisms linking autism and anxiety in monogenic cases of ID have not been delineated, and may be uncovered by future research investigating longitudinal relationships at the levels of neural systems, cognitive processes and interpersonal dynamics. One possibility is that structural and functional abnormalities in the amygdala and hippocampus, regions subserving emotion regulation which are also found to have decreased volumes in *DDX3X* and FXS mouse models (31, 32), represent a transdiagnostic risk mechanism underlying the co-occurrence of autism and anxiety in multiple monogenic disorders.

We also found that within the *DDX3X* group (only), anxiety predicted levels of SIBs. These findings may point to anxiety as a causal mechanism for SIBs in *DDX3X* variants, as has been demonstrated among autistic individuals without ID (33). However, a combination of individual, social and environmental factors are likely to underlie the occurrence of SIBs in people with ID (34), and interpreting underlying causes often requires reporters to make inferences about an individual’s internal state (35). Future research on the temporality of SIBs in *DDX3X* variants may clarify whether these behaviours are triggered by anxiety, or by other factors such as pain or frustration (36). At a mechanistic level, it is also important to situate SIBs in the context of the broader *DDX3X* phenotype. For example, it is possible that shared neural factors (e.g. those associated with emotion regulation) underlie social interactions, anxiety, impulsivity and sensory over- or under-reactivity, which converge to increase risk for multi-domain difficulties including SIBs. Interestingly, among girls and women with FXS, those who engage in SIBs are also more likely to have autism, anxiety and sensory processing issues (37). Further systematic investigation of the neural correlates, behavioural antecedents, and consequences of SIBs may clarify the aetiology and function of these behaviours in individuals with *DDX3X* variants, thus informing targeted prevention and response approaches (38).

Finally, we investigated the relevance of *DDX3X* variant type to anxiety and SIBs. In line with previous reports (9, 18), individuals with missense variants had poorer adaptive function compared to those with protein-truncating variants. Regression models revealed that variant type did not predict anxiety or SIBs. It is possible that genetic variant type may moderate developmental and behavioural factors contributing to these outcomes, but this could not be observed due to our small sample size and limited measures. On the other hand, true lack of association between variant type and social-emotional features may reflect the complex and multifactorial pathways influencing these outcomes, not predictable by genetic test result even in combination with other information. Future research should investigate the functional impact of specific variants – which may inform quantitative predictions of neurobiological impact – and also investigate non-genetic contributions to socioemotional outcomes.

### Strengths, limitations and future directions

Strengths of this study included the systematic post-diagnostic assessment of phenotypes in individuals diagnosed with *DDX3X* variants from multiple centres, and comparison to a mixed-aetiology ID group. However, we also recognise several key limitations to this work. First, we relied primarily on parent report questionnaires and interviews, which can be limited in scope, sensitivity and specificity. Future studies incorporating multi-informant perspectives, clinician-administered interview and observational schedules, and cognitive and physiological assessments that are tailored for individuals with ID will contribute richer insights into social and emotional functioning in individuals with *DDX3X* variants. Related to this, we did not examine how early and ongoing medical or physiological conditions (e.g. GI, sleep, motor issues) may impact developmental outcomes. This study included only girls and women with *DDX3X* variants, and future research should determine whether a similar profile of social and emotional experiences is present in males. In addition, this was a cross-sectional study, and participants’ ages spanned a broad range. Longitudinal studies delineating changes in adaptive, socioemotional and neuropsychological functioning will be crucial for mapping the developmental progression of *DDX3X*-associated difficulties and identifying periods and mechanisms of risk and resilience.

## Conclusions

*DDX3X* variants are increasingly recognised as a common cause of neurodevelopmental disorders in girls and women, with numbers of diagnosed individuals expected to rise as availability of testing increases. In this study, we expanded our knowledge of the social and emotional experiences of girls and young women with *DDX3X* variants, and highlighted a range of features that warrant clinical attention and further investigation. In common with other genomic disorders, it is important to place this diagnosis and its variable expression in the broader context of cognitive, psychological and interpersonal development. We find that rates of autistic characteristics do not differentiate *DDX3X* and ID-comparison groups, but anxiety and self-injury levels are higher in the *DDX3X* group. Autism and anxiety, and anxiety and self-injury, are closely linked within the *DDX3X* group and are not explained by global adaptive function. Future research investigating the diversity of mechanisms contributing to social and emotional outcomes will guide the development of more precise information and support available to individuals with *DDX3X* variants and their families, and contribute to a better understanding of autism and mental health amongst young women with ID more broadly.

## Data Availability

The data that support the findings of this study are available to other ethically approved research projects from the Principal Investigator, KB at.

## List of abbreviations

*DDX3X*: DEAD-box helicase 3 X-linked
*ID*: intellectual disability
*ADHD*: Attention Deficit / Hyperactivity Disorder
*MHI*: Medical History Interview
*VABS*: Vineland Adaptive Behaviour Scales
*SRS*: Social Responsiveness Scale, second edition
*DBC*: Developmental Behaviour Checklist, second edition
*ADI-R*: Autism Diagnostic Interview, Revised
*SIB*: Self injurious behaviour
*FXS*: Fragile X Syndrome

## Declarations

### Ethics approval and consent to participate

This study was approved by the Cambridge Central Research Ethics Committee (‘Phenotypes in Intellectual Disability’, reference: 11/EE/0330). Parents or Carers gave written informed consent on behalf of participants under the age of 16. For participants aged 16 and over, lacking capacity to consent, a consultee was appointed.

### Consent for publication

Obtained within study consent form.

## Authors’ contributions

ENC and SOB acquired the data. ENC analysed and interpreted the data, and drafted the paper. All other authors substantially revised it. DEA, GS, and KB conceived and designed the study. All authors read and approved the final manuscript.

## Acknowledgements

We sincerely thank the study participants, their families, and carers for their dedication and time in contributing to this study. We are grateful to the DDX3X Support UK group, the Deciphering Developmental Disorders (DDD) study, the IMAGINE-ID study, UK regional genetic centres, diagnostic laboratories, and referring clinicians for enabling us to publicise this study to the families who have taken part.

---

**Supplementary Table 1.**
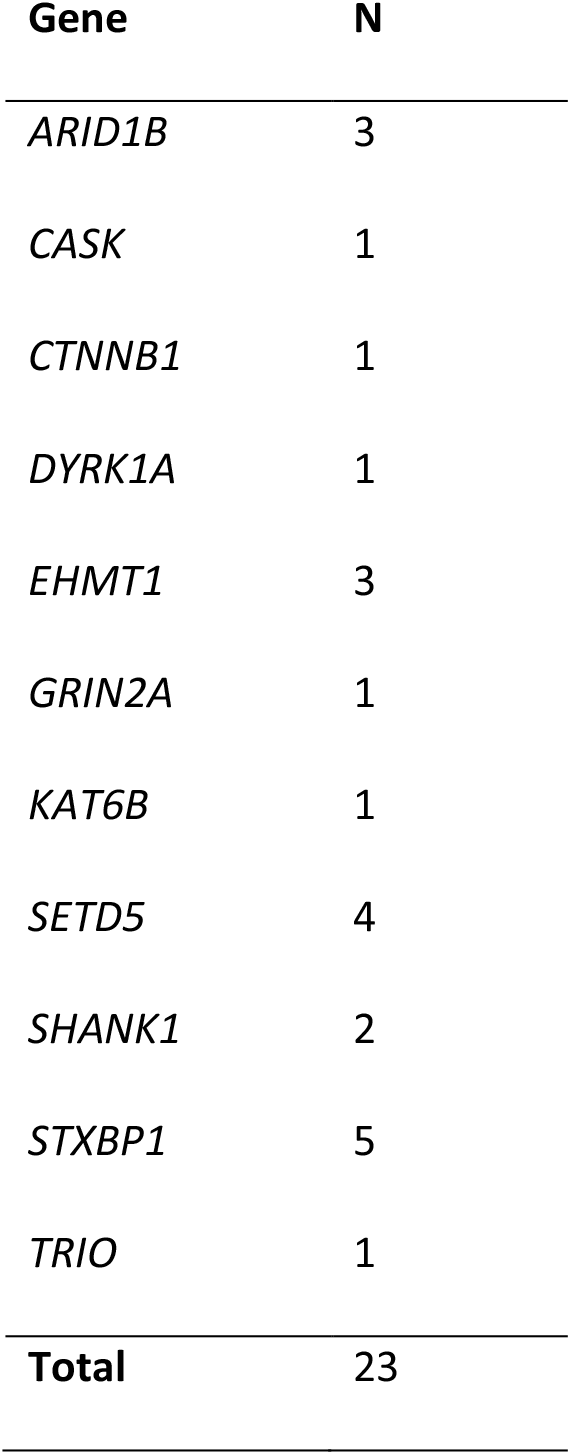
Genetic diagnoses in the ID-comparison group.

**Supplementary Table 2.** Participants’ gene variant information.

| Participant<br>no. | Decipher/100<br>K Genome ID | Inheritance | Variant type | Amino acid<br>change | Nucleotide<br>change | Pathogenicity | ID severity |
| --- | --- | --- | --- | --- | --- | --- | --- |
| 1 | 272869 | De novo | Missense | p.Cys467Tyr | c.1400G>A | Likely<br>pathogenic | Severe |
| 2 | 273466 | De novo | Splice<br>(acceptor) | - | c.46-2A>G | Likely<br>pathogenic | Mild |
| 3 | 272597 | De novo | Stop-gained | p.Ser24Ter | c.71C>A | Pathogenic | Mild |
| 4 | 264193 | De novo | Stop-gained | p.Arg291Ter | c.871C>T | Likely<br>pathogenic | Mild |
| 5 | 261890 | Inherited | Missense | p.Arg533His | c.1598G>A | Likely<br>pathogenic | Moderate |
| 6 | 267102 | De novo | Missense | p.Ala392Pro | c.1174G>C | Likely<br>pathogenic | Severe |
| 7 | 267105 | De novo | Missense | p.Ala392Pro | c.1174G>C | Likely<br>pathogenic | Moderate |
| 8 | 261162 | De novo | Stop-gained | p.Arg45Ter | c.133C>T | Pathogenic | Moderate |
| 9 | 290561 | De novo | Splice (donor) | - | c.281+1G>C | Likely<br>pathogenic | Moderate |
| 10 | 300283 | De novo | Missense | p.Arg474Cys | c.1420C>T | Likely<br>pathogenic | Mild |
| 11 | - | - | - | - | - | - | Moderate |
| 12 | 277307 | De novo | Splice (donor) | - | c.45+1G>T | Pathogenic | Mild |
| 13 | 303778 | De novo | Frameshift | p.Lys208Serfs<br>Ter13 | c.623delA | Pathogenic | Borderline |
| 14 | - | De novo | Missense | Cys341Arg | c.1021T>C | - | Severe |
| 15 | 282969 | De novo | Splice donor | - | c.45+1G>A | Pathogenic | Profound |
| 16 | - | - | - | - | - | - | Moderate |
| 17 | - | De novo | Splice<br>(acceptor) | - | c.766-8_766-2del | Likely<br>pathogenic | Mild |
| 18 | - | - | - | - | - | - | Moderate |
| 19 | - | Unknown | Frameshift | p.Asn124Lysfs<br>Ter5 | c.372_373del<br>CA | - | Mild |
| 20 | 307552 | De novo | Frameshift | p.Phe83Ter | c.241_242insC<br>TAG | Pathogenic | Borderline |
| 21 | 210015169 | De novo | Frameshift | p.Gly153Leufs<br>*69 | c.455_456dup<br>CT | Pathogenic | Mild |
| 22 | 281511 | De novo | Missense | p.Ser542Leu | c.1625C>T | Likely<br>pathogenic | Moderate |
| 23 | - | Unknown | Splice site | - | c.765+1G>A | Pathogenic | Mild |

**Supplementary Figure 1.**
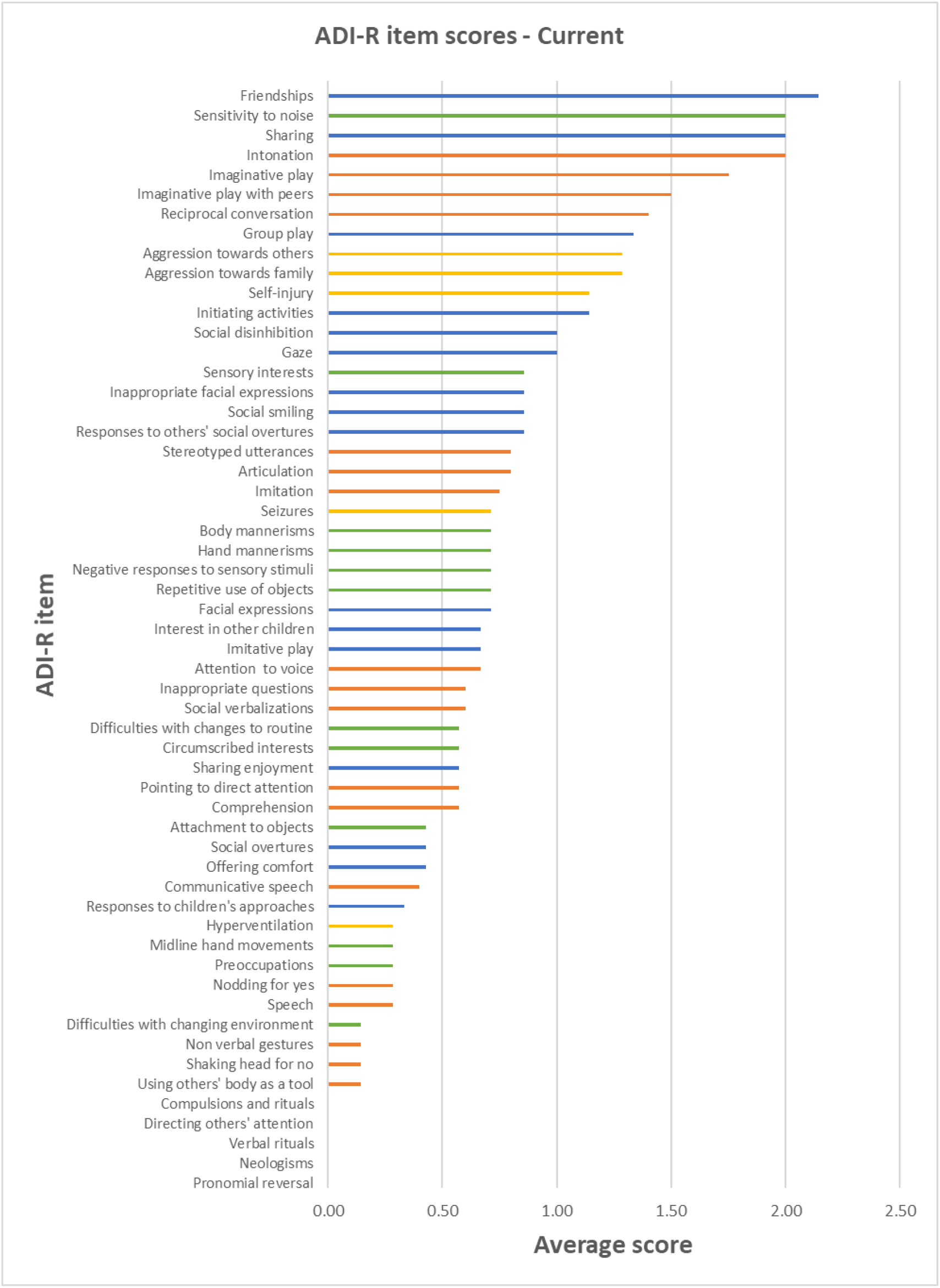
ADI score profiles – Current. *Note*. Orange items = Language and Communication, Blue = Social Development and Play, Green = Interests and Behaviours, Yellow = General Behaviours.

**Supplementary Figure 2.**
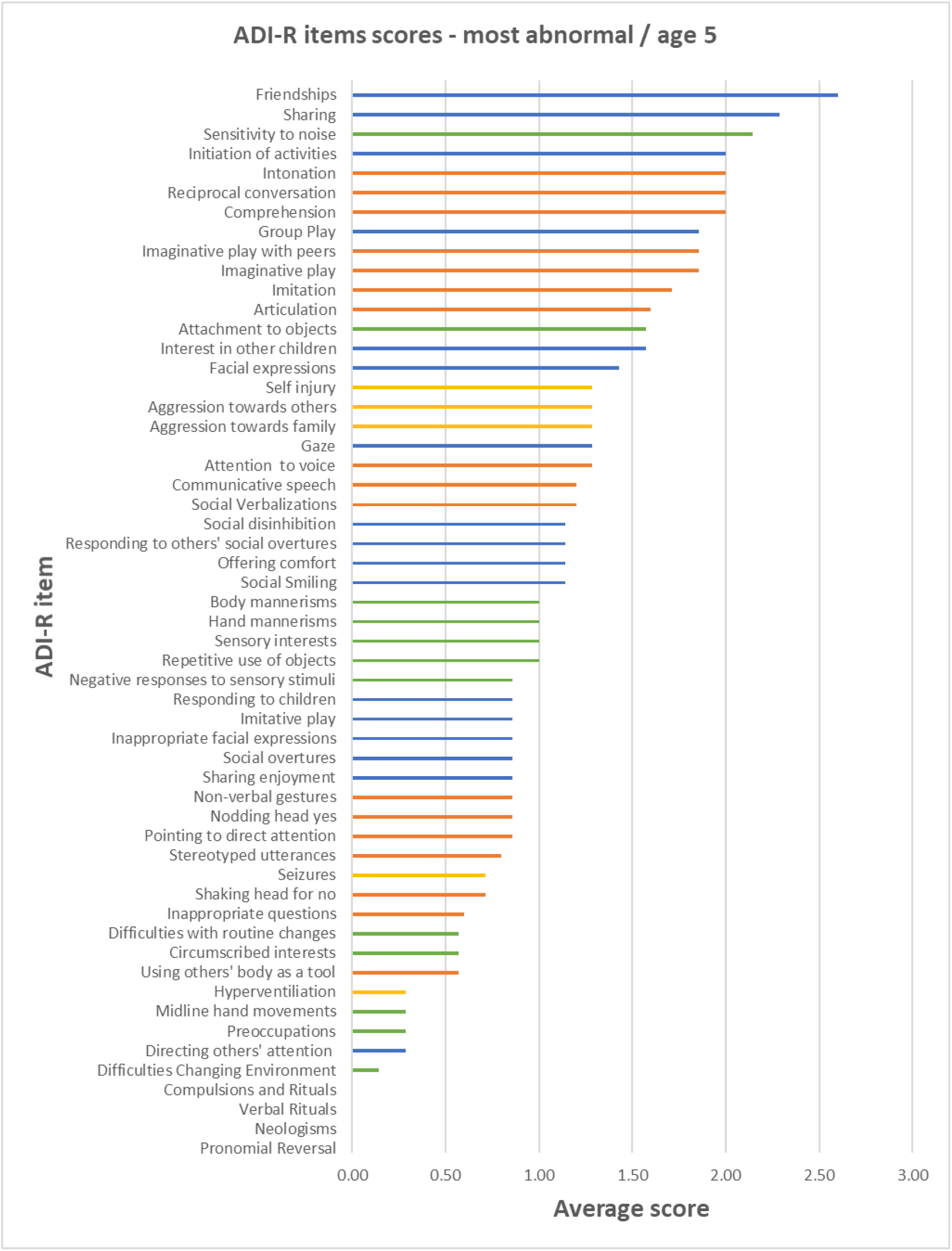
ADI score profiles – Most Abnormal / age 5. *Note*. Orange items = Language and Communication, Blue = Social Development and Play, Green = Interests and Behaviours, Yellow = General Behaviours.

